# Risk Factors and Clinical Presentations of Long COVID: A Retrospective Observational Propensity Score Matching Study

**DOI:** 10.1101/2025.03.01.25323136

**Authors:** Lanre Peter Daodu, Yogini Raste, Francesca I.F. Arrigoni, Judith E. Allgrove, Reem Kayyali

**Affiliations:** Faculty of Health, Sciences, Social Care and Education, Kingston University, London; Chest/Respiratory Department, Croydon University Hospital, London, Croydon Health Services NHS Trust

**Keywords:** COVID-19, long COVID, risk factors, symptoms, propensity score matching

## Abstract

**Introduction:** Long COVID symptoms can affect multiple body systems and result in complicated clinical presentations. Research on long COVID is fraught with various challenges, particularly identifying risk factors, understanding pathophysiology, and implementing effective management strategies. This study assessed long COVID risk factors and evaluated the symptoms presented by affected individuals.

**Methods:** We conducted a hospital-based single-centre retrospective study of 627 adult patients managed for COVID-19. The patients were divided into long COVID and non-long COVID cohorts. We employed propensity score matching (PSM) to create matched groups of participants based on their predicted risk of long COVID and subsequently calculated the multiplicative effects on the odds of developing it. The prevalence of each clinical symptom was calculated as a percentage to determine the most common.

**Results:** The study included 627 COVID-19 patients, with an average age of 59 (±20) years. The cohort included 375 without and 252 with long COVID. Each additional year of age increased the odds of long COVID by 6.4% (OR 1.06, p=0.00). BMI (OR 1.03, p=0.04), Asian or Asian British (OR 2.64, p = 0.03), and former smokers (OR 6.44, p=0.05) had significantly higher odds of long COVID. Long COVID risk rose with the number of comorbidities. Interstitial lung diseases (ILDs) (OR 2.93, p=0.02), high Clauss fibrinogen level (OR 1.22, p=0.00), and serum Sodium (Na^+^) (OR 1.07, p = 0.02) significantly increased the odds of long COVID. The most prevalent clinical presentations were fatigue (81%), shortness of breath (61%) and dry cough (19%).

**Conclusions:** The study identified key risk factors for long COVID, including age, BMI, ethnicity, former smoking habits, comorbidities, and ILDs. Clauss fibrinogen and serum Na^+^ were also significant biomarkers. The main symptoms were fatigue, shortness of breath, and dry cough. The multifaceted effects of long COVID necessitate a multidisciplinary approach to patient care.

**Key Messages:** *What is already known on this topic:*

- Advanced age, BMI, a history of severe COVID-19 infection, existing comorbidities, and chronic conditions such as diabetes mellitus (DM) were identified as risk factors for long COVID, and the symptoms frequently reported include fatigue, dyspnoea, cough, muscle and joint pain, cognitive deficits, and respiratory problems.

*What this study adds:*

- Ethnic minorities (Asian or Asian British), ILDs, elevated levels of Clauss fibrinogen and serum Na^+^ increased the odds of developing long COVID.

*How this study might affect research, practice, or policy:*

- The observed association between elevated Clauss fibrinogen levels and serum Na+ and long COVID necessitates in-depth research into the role of fibrinogen and deranged electrolytes in the pathophysiological processes of long COVID. Elucidating these mechanisms could facilitate the development of predictive models and targeted therapeutic interventions.

## INTRODUCTION

Long COVID has become a significant concern for clinicians, patients and the public. It has been described as *’pandemic after the pandemic’* ^1,2^. The widespread effects of the emerging long COVID phenomenon on millions of individuals worldwide highlight the importance of various research studies and their implications for global clinical practices and healthcare systems. The disease presents numerous research challenges and subjects of extensive research and debate, particularly in the areas of aetiology, risk factors, pathophysiology and efficacious treatments^3,4^.

Long COVID has been linked to several risk factors, with age consistently emerging as a significant determinant across diverse populations. Studies have shown that advanced age is a significant risk factor for long COVID in adults, with older individuals being more likely to experience prolonged symptoms ^5,6^. In addition, the severity of the initial COVID-19 illness has been correlated with a higher likelihood of developing long COVID. Patients who experienced more severe cases, as indicated by minimum oxygen saturation levels, were at a higher risk of developing long COVID ^7^. Also, pre-existing comorbidities have been recognised as a significant risk factor for long COVID. Chronic conditions, such as diabetes, have been linked to an increased risk, indicating that individuals with underlying health issues may be more susceptible to prolonged symptoms^8^.

The exact causes of the disease condition are not fully known. The disease involves a complex pathophysiology, including persistent immune activation, dysregulation, organ damage, microvascular dysfunction, and neurological changes^9^. It has been postulated that long COVID arises from tissue damage due to virus-specific changes or prolonged inflammatory responses triggered by viral persistence, immune dysregulation, and autoimmune reactions ^9, 10^. Dysregulated T-cell activation, chronic inflammation, oxidative stress, mitochondrial dysfunction, and endothelial dysfunction are key features of the pathophysiology of long COVID^11, 12^. Factors such as viral persistence, immune system abnormalities, dysregulated inflammatory responses, dysbiosis, reactivation of other viruses, and microthrombi formation further complicate the understanding of long COVID pathophysiology^13^. A recent study found that individuals with long COVID exhibit a specific blood protein signature characterised by heightened complement activation and thromboinflammation^14^. This includes activated platelets and indicators of red blood cell breakdown.

Studies have also highlighted the diverse spectrum of symptoms and signs associated with long COVID. Symptoms commonly reported include fatigue, dyspnoea, cough, muscle and joint aches and cognitive deficits^15-19^. Additionally, gastrointestinal symptoms have been linked to specific bacterial colonisation and alterations in serum metabolites^20, 21^. Studies have attempted to categorise symptoms into core and extended clusters based on disease domains to facilitate clinical assessment and research^22^. The impacts of persistent symptoms in individuals with long COVID-19 are substantial. Individuals with long COVID may experience functional limitations, which impact their housing security and lead to challenges in work productivity and health-related quality of life^23,24^. The persistence of symptoms like fatigue, musculoskeletal pain, cardiothoracic symptoms, and neurocognitive impairment can significantly impair daily functioning and well-being^25^.

Despite substantial progress in understanding long COVID, the risk factors contributing to the development of this condition and its diverse clinical presentations remain poorly defined. The heterogeneity in symptom presentation complicates the identification of risk factors and the development of targeted treatments ^26,27^. Addressing this gap is essential for identifying individuals at higher risk of developing long COVID. This knowledge can inform clinical practices to mitigate the impact of long COVID. Additionally, there are disparities in the prevalence and severity of long COVID among different ethnic and racial groups^28, 29^. Research in this area can lead to more inclusive and representative studies, providing an understanding of long COVID among a diverse population. Also, few studies have used propensity score matching to assess the risk factors for COVID. This methodological gap is essential as propensity score matching (PSM) can help reduce bias and confounding variables, leading to more accurate and reliable results^30^.

This study assessed the potential risk factors associated with the likelihood of experiencing long COVID symptoms and analysed the symptoms exhibited by individuals affected by this chronic condition. The study addressed the following primary research question: What are the potential risk factors linked to the likelihood of experiencing long COVID symptoms in individuals who have recuperated from SARS-CoV-2 infection?

This retrospective observational study employed PSM to compare individuals who developed long COVID with those who did not, based on patients’ demographics, clinical characteristics, comorbidities, vaccination history and laboratory biomarkers obtained from electronic patient records (EPRs). PSM minimised the bias^31^ that might arise from differences in baseline characteristics, allowing for a more accurate assessment of risk factors and symptomology.

## MATERIALS AND METHODS

### Study Design and Setting

A retrospective observational study was conducted from June 2023 to June 2024 at the Croydon University Hospital (CUH) in South London, England. The study aimed to anonymously review the EPRs of 627 patients who were managed for COVID-19 between April 2020 and December 2022.

The study was limited to using previously collected, non-identifiable information. There was no contact or follow-up with patients for this study.

Findings were reported in accordance with the Strengthening the Reporting of Observational Studies in Epidemiology (STROBE) guidelines^32^.

### Participants

The study included adult patients aged 18 years or older, consisting of 627 individuals who tested positive for and were managed for COVID-19. The exclusion criteria involved paediatric patients confirmed positive for COVID-19 and those lacking COVID-19 test results. Additionally, we excluded all patients who were confirmed as deceased during the acute phase of the SARS-CoV-2 virus. Test results were used to identify all individuals with COVID-19, while physicians’ clinical assessment and diagnosis were used to determine individuals with long COVID.

### Patient and Public Involvement

This was inapplicable due to the retrospective nature of the study.

### Data Collection

Data were entered into Castor Electronic Data Capture (EDC) software^33^. The collected data includes patients’ demographics, such as age, gender, and ethnicity. It also covered health indicators like smoking status, body mass index (BMI), number of comorbidities by each patient, and type of comorbidities: hypertension (HTN), other heart diseases, diabetes mellitus (DM), asthma, chronic obstructive pulmonary disease (COPD), interstitial lung diseases (ILDs), neurological diseases, chronic kidney disease (CKD) and cancers. COVID reinfection, vaccination history and laboratory biomarkers: Mean Corpuscular Haemoglobin Concentration (MCHC), Red Cell Distribution Width (RDW), International Normalized Ratio (INR), Activated Partial Thromboplastin Time (APTT), Clauss Fibrinogen, D-dimer, Alkaline Phosphatase (ALP), Alanine Aminotransferase (ALA), Total Bilirubin (TBIL), Creatine Kinase (CK), Albumin (ALB), Potassium (K+), Creatinine (Cr), Troponin T (TnT), Sodium (Na+), and Urea (Ur).

### Outcomes

The primary outcome was to identify potential risk factors associated with the likelihood of experiencing long COVID symptoms following recovery from SARS-CoV-2 infection. The secondary outcome focused on examining the clinical presentations of long-term COVID-19 in individuals who have recovered from the SARS-CoV-2 virus.

### Statistical Analysis

The statistical analysis was conducted using R programming version 4.3.3. Initially, a thorough approach was taken to address missing data in the covariates. The Visualization and Imputation of Missing Values (VIM) method was employed to visualise and understand the missing data patterns within the dataset^34^. Multiple imputations by chained equations (MICE) was used to impute the missing covariate data before estimating the propensity scores^35^. MICE was selected for its capability to impute missing data without requiring the specification of a joint distribution for the covariates^36^.

Descriptive statistics were used to analyse and communicate the fundamental attributes of the data through suitable statistical tests. Mean and standard deviation (SD) were used for continuous variables, and counts and percentages were used for categorical variables. The Wilcoxon rank sum test was used to compare continuous variables between the two cohorts. Pearson’s Chi-squared test was applied to categorical variables; the value was set at < 0.05 for significance.

### Propensity Score Matching (PSM)

We employed full matching (FM) to match all study participants based on their probability of developing long COVID. FM aligned long COVID and non-long COVID patients as closely as possible based on their propensity scores, estimated using a generalised linear model (GLM) with a probit link function. A calliper of 0.5 was applied, restricting the maximum allowable difference in propensity scores between matched units to improve the quality of the matches. FM (also called Optimal full matching) is particularly effective when there are multiple control units for each treated unit or when there is a significant difference in the distribution of propensity scores between the groups^37,38^. This form of matching minimises discrepancies between treated (long COVID group) and control (non-long COVID) units by forming matched sets, thereby maximising the use of available data and ensuring similarity in propensity scores^38^.

The covariates in the PSM included age, gender, ethnicity, BMI, smoking status, number of comorbidities by each patient, type of comorbidities: HTN, other heart diseases, DM, asthma and COPD, ILDs, neurological diseases, CKD and cancer, vaccination status, COVID-19 reinfection and Laboratory biomarkers: MCHC, RDW, INR, APTT, Clauss fibrinogen, D-dimer, ALP, ALA, TBIL, CK, ALB, K+, Cr, TnT, Na+ and Ur.

The matched data were extracted for further analysis by extracting matched datasets post-matching, commonly employed for propensity score matching in observational studies^37^. This step retrieved the subset of the original dataset that includes only the matched pairs, ensuring the analysis was based on a balanced sample where the long COVID and non-long COVID patients were comparable. Propensity scores were then extracted from the ’distance’ variable in the ’matched data’ frame and stored. These scores represent the likelihood of an individual possessing a particular characteristic or belonging to a specific group based on a set of observed covariates. Subsequently, these scores were matched to individuals to balance the distribution of observed covariates between groups.

Then, we employed a logistic regression model using the matched data to assess the relationship between the variables and long COVID. The logistic regression model was specified with a quasibinomial family to account for overdispersion in the binary outcome variable. We performed a multivariate analysis to interpret the logistic regression results by calculating the odds ratios and confidence intervals. By exponentiating the coefficients, we transformed the log-odds outputs into odds ratios, which were more interpretable as they represented the change in odds of developing long COVID occurring for a one-unit change in the predictor variable. These steps ensured a thorough and robust statistical analysis, enabling accurate interpretation of the relationships between the variables and the likelihood of experiencing long COVID symptoms.

### Clinical Presentation of Symptoms Estimation

The clinical presentation of symptoms among study participants with long COVID was quantified using percentages. The visualisation technique ^41^ was employed to identify the most prevalent symptoms and group each symptom to the body system: neurological, psychological, respiratory, cardiac, musculoskeletal, endocrine, gastrointestinal (GI) and immune systems.

## RESULTS

### Characteristics of the Study Patients

The total number of study participants was 627, with 375 classified as non-long COVID and 252 as long COVID patients (Table 1). Long COVID patients (mean age 69) were significantly older than non-long COVID patients (mean age 53, p = <0.00). There was no significant gender difference between groups (p = 0.30). Most patients (57% non-long COVID, 52% long COVID) were female. BMI was similar in both groups (avg 29-30, p=0.30). Groups showed similar ethnic distributions (p = 0.80). Most patients (41%) were White. Patient comorbidities varied significantly (p = <0.00). Long COVID patients showed a much higher rate of multiple comorbidities (35% vs 19%). Long COVID patients had more HTN (47% vs 27%, p = <0.00), other heart diseases (23% vs 16%, p = 0.02), DM (29% vs 19%, p=0.01), asthma/COPD (19% vs 13%, p = 0.03), ILDs (9.5% vs 2.7%, p = <0.00), neurological diseases (24% vs 21%, p = 0.05), CKD (8.3% vs 4.3%, p = 0.04) and cancer (9.1% vs 7.2%, p = 0.50). Long COVID patients had more former smokers (15% vs 11%, p=0.03), while non-long COVID had more current smokers (7.1% vs 9.9%). More patients in long COVID groups had COVID reinfection (5.2% vs 4.5%, p=0.70). Lab biomarker values differed significantly. Group differences in INR, fibrinogen, and creatinine were significant (p = <0.00). A significantly higher percentage of Long COVID patients (59%) received full vaccination plus booster doses than non-Long COVID patients (38%, p = <0.00).

**Table 1.** Characteristics of the Study 627 Patients

| Variable | Overall<br>N = 627 | Non-Long COVID<br>N=375 | Long COVID<br>N=252 | p-value<sup>3</sup> |
| --- | --- | --- | --- | --- |
| <sup>1</sup>Age (Year) | 59 (20) | 53 (20) | 69 (16) | <0.00 |
| <sup>2</sup>Gender (n (%)) |  |  |  | 0.30 |
| Female | 344(55%) | 213 (57%) | 131(52%) |  |
| Male | 283 (45%) | 162 (43%) | 121(48%) |  |
| <sup>1</sup>BMI | 29 (7) | 29 (7) | 30 (8) | 0.30 |
| <sup>2</sup>Ethnicity |  |  |  | 0.80 |
| Any other ethnic group | 90(14%) | 57(15%) | 33 (13%) |  |
| Asian/Asian British | 118(19%) | 65 (17%) | 53(21%) |  |
| Black, Black British, Caribbean/African | 145(23%) | 86(23%) | 59 (23%) |  |
| Mixed or multiple ethnic | 18(2.9%) | 12(3.2%) | 6 (2.4%) |  |
| White | 256(41%) | 155(41%) | 101(40%) |  |
| **<sup>2</sup>Number of comorbidities** |  |  |  | <0.00 |
| 1 | 95(15%) | 53 (14%) | 42(17%) |  |
| 2 | 85 (14%) | 44 (12%) | 41(16%) |  |
| 3 | 68(11%) | 37 (9.9%) | 31(12%) |  |
| 4 | 52(8.3%) | 25 (6.7%) | 27(11%) |  |
| 5 | 60 (9.6%) | 24(6.4%) | 36(14%) |  |
| ≥6 | 98(16%) | 46(12%) | 52(21%) |  |
| **<sup>2</sup>Types of comorbidities** |  |  |  |  |
| HTN | 221(35%) | 103(27%) | 118(47%) | <0.00 |
| Other Heart Diseases. | 118(19%) | 59 (16%) | 59(23%) | 0.02 |
| DM | 143(23%) | 71(19%) | 72(29%) | 0.01 |
| Asthma/COPD | 95(15%) | 47 (13%) | 48(19%) | 0.03 |
| ILDs | 34 (5.4%) | 10 (2.7%) | 24(9.5%) | <0.00 |
| Neurological Diseases | 140(22%) | 80 (21%) | 60 (24%) | 0.50 |
| CKD | 37 (5.9%) | 16(4.3%) | 21(8.3%) | 0.04 |
| Cancer | 50 (8.0%) | 27(7.2%) | 23 (9.1%) | 0.50 |
| **<sup>2</sup>Smoking Status** |  |  |  | 0.03 |
| Smoking (Current) | 55 (8.8%) | 37(9.9%) | 18(7.1%) |  |
| Smoking (Former) | 80(13%) | 42(11%) | 38(15%) |  |
| **<sup>2</sup>COVID Reinfection** | 30(4.8%) | 17(4.5%) | 13(5.2%) | 0.70 |
| **<sup>\*1</sup>Laboratory Biomarker** |  |  |  |  |
| MCHC | 334 (11) | 334 (10) | 333 (11) | 0.70 |
| RDW | 14.98(9.89) | 15.32 (12.95) | 14.53 (1.80) | 0.80 |
| INR | 1.16 (0.70) | 1.15 (0.87) | 1.17 (0.39) | <0.00 |
| APTT ratio | 1.16 (3.87) | 1.30 (5.15) | 0.99 (0.15) | >0.90 |
| Clauss fibrinogen level | 5.63 (2.02) | 5.35 (2.01) | 5.99 (1.98) | <0.00 |
| D-dimer | 1,041(4,227) | 1,121 (5,219) | 957 (2,872) | >0.90 |
| ALP | 99 (62) | 102 (64) | 94 (60) | 0.20 |
| ALA | 47 (87) | 48 (105) | 47 (55) | 0.14 |
| TBIL | 9.4 (7.0) | 9.3 (7.0) | 9.4 (7.0) | 0.40 |
| CK | 259 (390) | 290 (436) | 224 (336) | >0.90 |
| ALB | 32.0 (5.4) | 32.5 (5.7) | 31.4 (4.9) | 0.02 |
| K+ | 4.48 (5.91) | 4.21 (0.48) | 4.81 (8.83) | 0.50 |
| Cr | 96 (102) | 85 (66) | 111 (132) | 0.00 |
| TnT | 28 (128) | 19 (26) | 36 (176) | 0.50 |
| Na+ | 137.0 (3.9) | 136.8 (3.6) | 137.3 (4.2) | 0.06 |
| Ur | 6.3 (5.0) | 5.6 (3.7) | 7.2 (6.1) | <0.00 |
| **<sup>2</sup>Vaccination Status** |  |  |  | <0.00 |
| *Fully vaccinated | 82(13%) | 52 (14%) | 30(12%) |  |
| Fully vaccinated plus booster doses | 293(47%) | 144(38%) | 149 (59%) |  |
| No vaccination | 234(37%) | 165 (44%) | 69 (27%) |  |
| *Partially vaccinated | 18 (2.9%) | 14(3.7%) | 4 (1.6%) |  |
| <sup>1</sup> Mean (SD) |  |  |  |  |
| <sup>2</sup> n (%) |  |  |  |  |
| <sup>3</sup> Wilcoxon rank sum test; Pearson's Chi-squared test |  |  |  |  |

\*Fully vaccinated: received the 2-dose series (Pfizer-BioNTech or Moderna or other vaccine authorised by WHO; \*Partial Vaccinated: received only one dose).
\*Laboratory biomarkers reference values : MCHC: 310-360g/L; RDW: 11.5-15%; D-dimer: 21-300ng/L; INR: 0.8-1.1; APTT: 0.85-1.15; Clauss Fibrinogen: 1.6-4.8g/L; TnT: 0-14ng/L; ALB: 35-50g/L; Serum Cr: 60-106umol/L; Serum K+: 3.5-5.3mmol/L; Serum Na+: 136-146mmol/L; Serum Ur: 2.5 – 7.8mmol/L and CK : (Croydon University Hospital London, reference values).

## Propensity Score Matching

### Covariate Balance Before and After Matching

Figure 1 and Table 2 show the standardised mean differences (SMDs) of covariates before and after matching. The vertical dashed lines indicated thresholds for acceptable balance, typically within -0.1 to 0.1 (Figure 1). Before matching, many covariates SMDs indicated significant imbalances except for covariates such as male, Asian or Asian British, three comorbidities, TnT, COVID reinfection, INR, cancer, one comorbidity, White, BMI, smoking(Former), Mixed or multiple ethnic groups, Fully vaccinated, D-dimer, Smoking(Current), neurological diseases, CK, Black, Black British, Caribbean or African, MCHC, any other ethnic group and female were within the acceptable balance thresholds.

**Figure 1.**
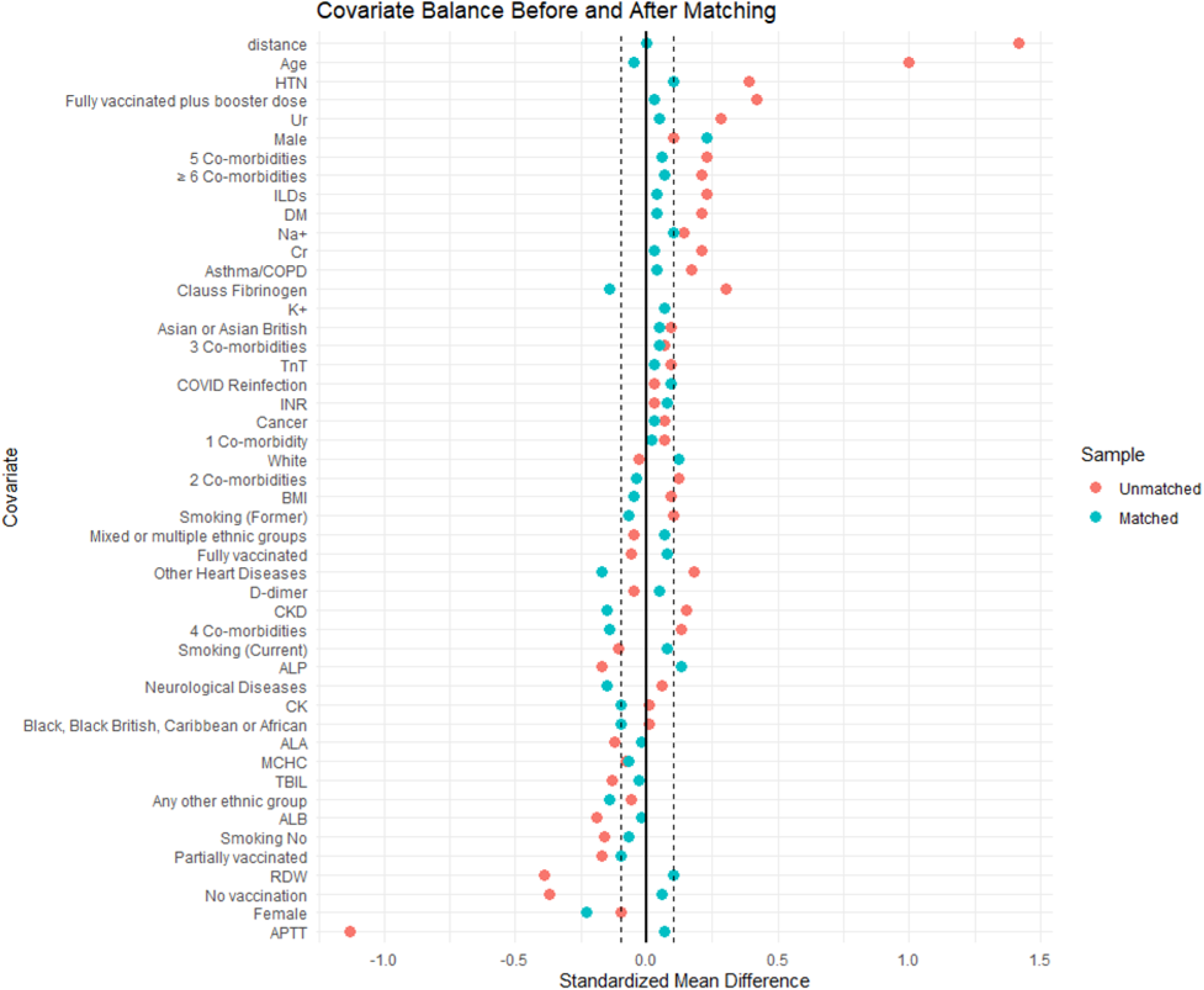
A Love plot to assess the balance of covariates after matching

**Table 2.** Standardised Mean Differences (SMD) of Covariates Before and After Matching

| Covariate | SMD Before Matching | SMD After Matching |
| --- | --- | --- |
| distance | 1.42 | 0.00 |
| Age | 1.0 | -0.05 |
| Female | -0.10 | -0.23 |
| Male | 0.10 | 0.23 |
| BMI | 0.09 | -0.05 |
| Any other ethnic group | -0.06 | -0.14 |
| Asian or Asian British | 0.09 | 0.05 |
| Black, Black British, Caribbean or African | 0.01 | -0.10 |
| Mixed or multiple ethnic groups | -0.05 | 0.07 |
| White | -0.03 | 0.12 |
| Smoking No | -0.16 | -0.07 |
| Smoking (Current) | -0.11 | 0.08 |
| Smoking (Former) | 0.10 | -0.07 |
| HTN | 0.39 | 0.10 |
| 1 Co-morbidity | 0.07 | 0.02 |
| 2 Co-morbidities | 0.12 | -0.04 |
| 3 Co-morbidities | 0.07 | 0.05 |
| 4 Co-morbidities | 0.13 | -0.14 |
| 5 Co-morbidities | 0.23 | 0.06 |
| ≥ 6 Co-morbidities | 0.21 | 0.07 |
| Other Heart Diseases | 0.18 | -0.17 |
| DM | 0.21 | 0.04 |
| Asthma/COPD | 0.17 | 0.04 |
| ILDs | 0.23 | 0.04 |
| Neurological Diseases | 0.06 | -0.15 |
| CKD | 0.15 | -0.15 |
| Cancer | 0.07 | 0.03 |
| COVID Reinfection | 0.03 | 0.09 |
| MCHC | -0.08 | -0.07 |
| RDW | -0.39 | 0.10 |
| INR | 0.03 | 0.08 |
| APTT | -1.13 | 0.07 |
| Clauss Fibrinogen | 0.30 | -0.14 |
| D-dimer | -0.05 | 0.05 |
| ALP | -0.17 | 0.13 |
| ALA | -0.12 | -0.02 |
| TBIL | -0.13 | -0.03 |
| CK | 0.01 | -0.10 |
| ALB | -0.19 | -0.02 |
| K+ | 0.07 | 0.07 |
| Cr | 0.21 | 0.03 |
| TnT | 0.09 | 0.03 |
| Na+ | 0.14 | 0.10 |
| Ur | 0.28 | 0.05 |
| Fully vaccinated | -0.06 | 0.08 |
| Fully vaccinated plus booster dose | 0.42 | 0.03 |
| No vaccination | -0.37 | -0.06 |
| Partially vaccinated | -0.17 | -0.10 |

After matching, the SMD values were significantly reduced, indicating improved group balance within the acceptable balance thresholds except for covariates such as Clauss fibrinogen, ALP, other heart diseases, CKD, four co-comorbidities, neurological diseases, any other ethnic groups, female, male and White.

## Propensity score distribution between the two groups

The y-axis measured the propensity scores ranging from 0.00 to 1.20 (Figure 2). Each group was illustrated by a combination of a violin plot, a box plot, and a mean point. The violin plot showed the density of the propensity scores at different values. The width of the plot at any given y-value reflected the number of data points at that score level, providing a visual depiction of data concentration. A box plot was embedded within each violin plot, showing the median propensity score as a central line, with the interquartile range (IQR) indicated by the box. The whiskers extended to the most extreme data points within 1.5 times the IQR. The presence of outliers was confirmed, as the box plots displayed points beyond the whiskers. The black diamond shapes represented the mean propensity scores for each group. For the non-long COVID patients, the propensity scores were densely concentrated around a median score of approximately 0.25, with most scores falling within the IQR of approximately 0.1 to 0.50. This concentration around the median indicated a central tendency in this group’s propensity scores. The long COVID group, however, showed a propensity score distribution centred around a median score close to 0.60, with an IQR of about 0.50 to 0.80. The density of scores in this group was more evenly spread across the range, suggesting a more uniform distribution than the non-long COVID group.

**Figure 2.**
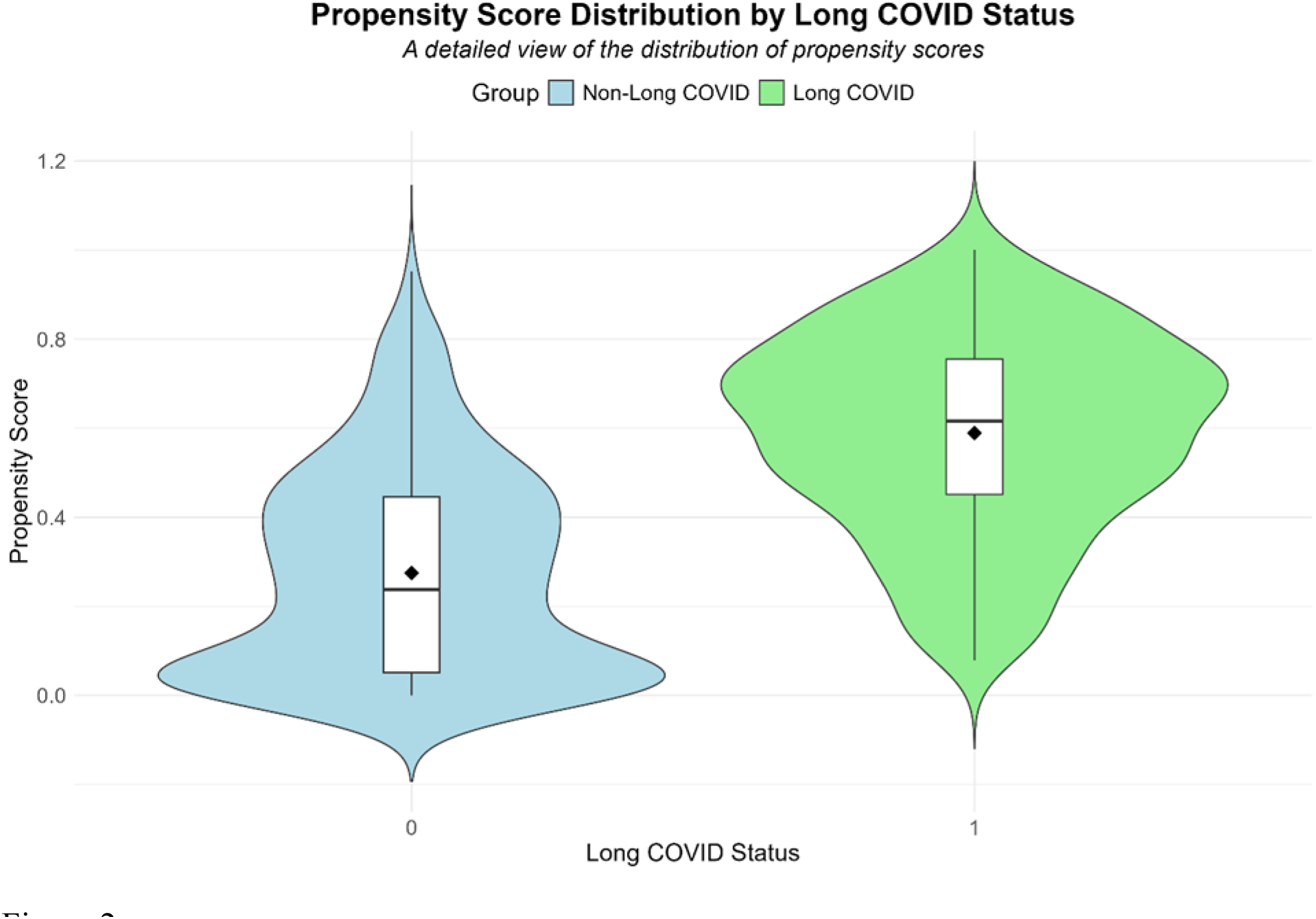
A violin plot that displays the propensity score distribution for long COVID and non-Long COVID

## The potential risk factors and the likelihood of developing long COVID

Table 3 shows a significant intercept (OR 0.00, p = 0.02). This indicated the baseline log-odds of the developing long COVID when all predictors were zero. The strength and direction of the association between each variable and the long COVID were shown by the OR. OR > 1 suggested a higher probability of the outcome, while values < 1 suggested a lower probability, and OR = 1 suggested no association.

**Table 3.** Multiplicative effects on the odds of long COVID

| Variable | OR | CI, 95% | p-Value |
| --- | --- | --- | --- |
| (Intercept) | 0.00 | 0.00 - 0.13 | 0.02 |
| Age | 1.06 | 1.02 - 1.11 | 0.00 |
| Male | 1.33 | 0.81 - 2.19 | 0.26 |
| BMI | 1.03 | 1.00 - 1.06 | 0.04 |
| Asian or Asian British | 2.64 | 1.11 - 6.41 | 0.03 |
| Black, Black British, Caribbean/African | 1.45 | 0.63 - 3.39 | 0.39 |
| Mixed or multiple ethnic | 4.25 | 0.73 – 23.07 | 0.10 |
| White | 1.29 | 0.57 – 2.96 | 0.54 |
| Smoking (Current) | 2.19 | 0.22 – 16.65 | 0.46 |
| Smoking (Former) | 6.44 | 1.14 – 54.16 | 0.05 |
| 1 Comorbidity | 3.87 | 1.88 – 8.14 | 0.00 |
| 2 Comorbidities | 2.96 | 1.38 - 6.48 | 0.01 |
| 3 Comorbidities | 2.16 | 0.94 – 5.02 | 0.07 |
| 4 Comorbidities | 3.25 | 1.32 - 8.09 | 0.01 |
| 5 Comorbidities | 4.13 | 1.62 – 10.76 | 0.00 |
| ≥ 6 Comorbidities | 2.79 | 1.17 – 6.71 | 0.02 |
| HTN | 1.11 | 0.68 - 1.81 | 0.67 |
| Other Heart Diseases | 0.67 | 0.38 - 1.18 | 0.17 |
| DM | 0.75 | 0.44 - 1.26 | 0.28 |
| Asthma/COPD | 2.15 | 0.98 - 4.83 | 0.06 |
| ILDs | 2.93 | 1.23 - 7.48 | 0.02 |
| Neurological Diseases | 0.63 | 0.38 - 1.04 | 0.07 |
| CKD | 1.58 | 0.66 - 3.87 | 0.31 |
| Cancer | 0.71 | 0.34 - 1.45 | 0.35 |
| Reinfection | 2.00 | 0.68 – 5.78 | 0.20 |
| MCHC | 1.00 | 0.98 - 1.02 | 1.00 |
| RDW | 0.98 | 0.86 - 1.01 | 0.55 |
| INR | 1.10 | 0.78 - 1.48 | 0.57 |
| APTT | 0.99 | NA - 1.06 | 0.88 |
| Clauss Fibrinogen | 1.22 | 1.08 - 1.38 | 0.00 |
| D-dimer | 1.00 | 1.00 - 1.00 | 0.17 |
| ALP | 1.00 | 0.99 - 1.00 | 0.14 |
| ALA | 1.00 | 1.00 - 1.00 | 0.46 |
| TBIL | 0.97 | 0.94 - 1.01 | 0.18 |
| CK | 1.00 | 1.00 - 1.00 | 0.05 |
| ALB | 0.97 | 0.92 - 1.02 | 0.24 |
| K+ | 1.03 | 0.98 - NA | 0.59 |
| Cr | 1.00 | 1.00 - 1.01 | 0.03 |
| TnT | 1.00 | 1.00 - 1.00 | 0.24 |
| Na+ | 1.07 | 1.01 - 1.14 | 0.02 |
| Ur | 0.99 | 0.92 - 1.05 | 0.70 |
| Fully vaccinated plus booster doses | 6.49 | 0.65 – 8.15 | 0.13 |
| No vaccination | 0.31 | 0.03 – 4.45 | 0.37 |
| Partially vaccinated | 2.08 | 0.01 – 134.86 | 0.75 |

Age was a significant predictor of long COVID (OR 1.06, p = 0.00). Each additional year of age increased the odds of developing long COVID by 6.4%. Males showed a non-significant increase in odds (OR 1.21, p=0.47). The odds of long COVID were slightly higher with increased BMI (OR 1.03, p=0.04), indicating a statistically significant relationship. Asian or Asian British (OR 2.64, p = 0.03) had more than twice the risk of experiencing long COVID, which was statistically significant. However, mixed or multiple ethnicities had a strong association (OR 4.25, p=0.10), Black, British, Caribbean, or African individuals had a moderate association (OR 1.45, p=0.39), and White individuals had a low association (OR 1.29, p=0.54); none of them was significant.

Former smokers (OR 6.44, p = 0.05) had significantly higher odds of developing long COVID, while current smokers (OR 2.19, p = 0.46) did not show a significant association. A significant association was found between the number of comorbidities and long COVID risk; odds ratios increased with each additional comorbidity except for patients with three comorbidities (one: OR 3.87, p=0.00; two: OR 2.96, p=0.01; three: OR 2.16, p=0.07; four: OR 3.25, p=0.01; five: OR 4.13, p=0.00, ≥ six: OR 2.79, p=0.02).

ILDs (OR 2.93, p = 0.02) increased the odds of developing long COVID, which was statistically significant, while other heart diseases (OR 0.67, p=0.67), HTN (OR 1.11, p =0.67), DM (OR 0.75, p=0.28), Asthma/COPD (OR 2.15, p = 0.06), neurological diseases (OR 0.63, p = 0.07), CKD (OR 1.58, p=0.31) and cancer (OR 0.71, p=0.35) were not significantly associated with the odds of developing long COVID. COVID reinfection (OR 2.00, p = 0.74) showed no significant association.

A significant association was found between elevated Clauss fibrinogen levels and the probability of long COVID (OR 1.21, p=0.00). In addition, a significant association was observed between elevated serum sodium levels and an increased probability of developing long COVID (OR 1.07, p = 0.02). There was no significant association between Cr levels (OR 1.00, p=0.03) and CK levels (OR 1.00, p=0.05). Other markers like MCHC, RDW, INR D-dimer, ALP, ALA, TBIL, ALB, K+, TnT, and Ur showed non-significant associations.

Fully vaccinated plus booster doses (OR 6.49, p=0.13), No vaccination (OR 0.31, p = 0.37), and partially vaccinated (OR 2.08, p = 0.75) showed non-significant associations.

## Clinical presentations of long COVID patients

A study of 252 patients with long COVID revealed a wide range of symptoms affecting multiple body systems, as depicted in Figure 3. The most common symptoms were fatigue (81%), shortness of breath (61%), dry/nonproductive cough (19%), chest pain (12%), short-term memory issues (12%), other memory difficulties (12%), anxiety (9.1%), arthralgia (6.8%), chest tightness (6.7%), cough with phlegm/productive cough (6.3%), depression (4.8%), poor mobility (4.8%), myalgia (4.8%), back pain (4%), ageusia (2.4%), poor balance (2.4%), dizziness and /lightheadedness (2.4%). Others include headaches (1.6%), difficulty in organising and planning (1.6%), anosmia (1.6%), hair fall, hair loss, and lethargy(1.6%), headaches (1.6%), confusion (1.2%), palpitations (1.2%), agoraphobia (0.8%), and anorexia (0.8%), tingling, sleep inertia, peripheral numbness and hypersomnia. (0.8%), diarrhoea 0.8%, rhinorrhoea (0.4%) and fever(0.4%). Most symptoms were associated with the neurological system, followed by the psychological, respiratory, musculoskeletal, cardiac, endocrine, and GI systems were less common.

**Figure 3.**
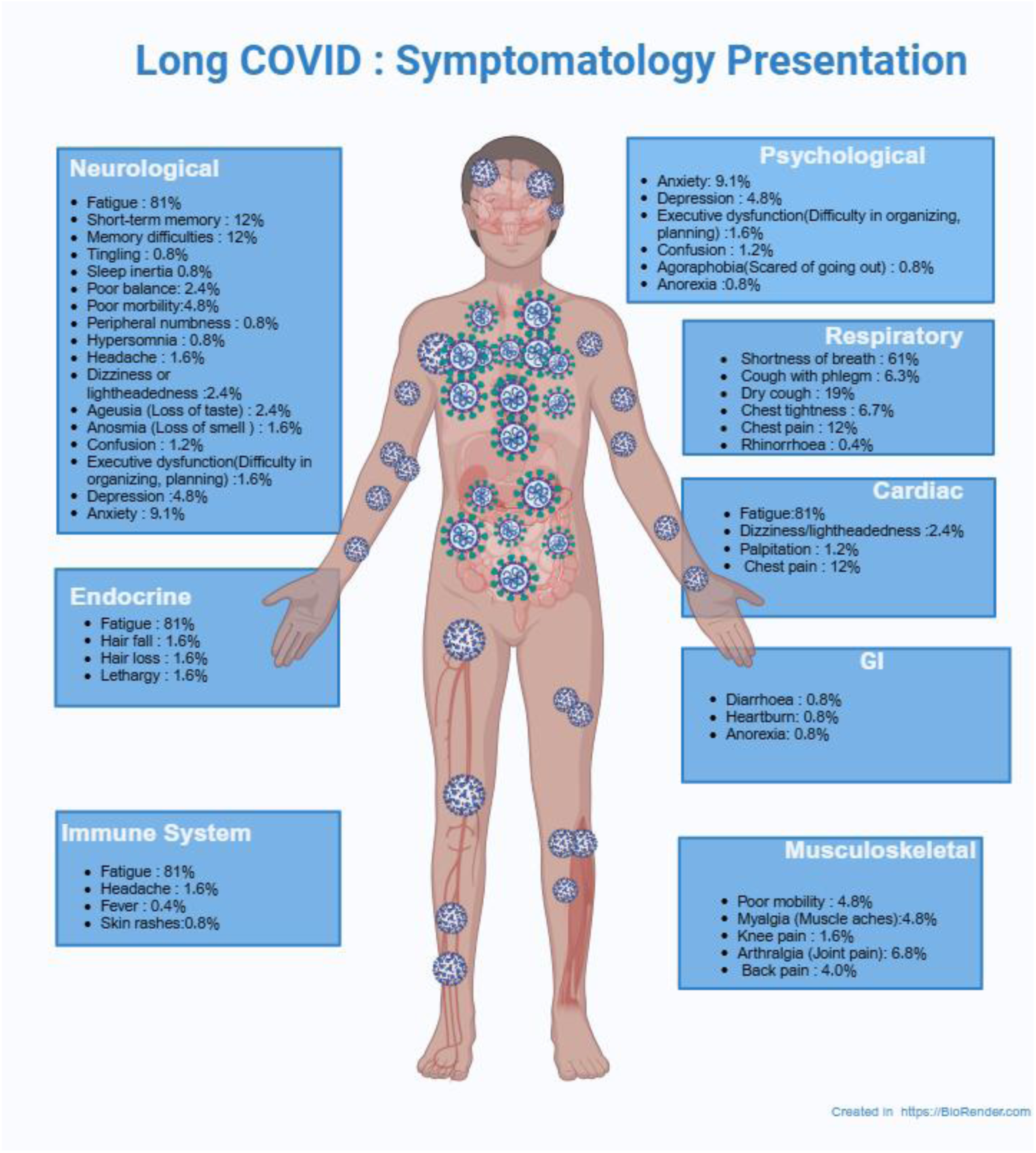
A wide range of long COVID symptoms affecting multiple body systems

## Discussion

### Summary of Keys Findings

Age was identified as a significant predictor, with every additional year increasing the likelihood of experiencing long COVID by 6.4%. Also, a higher BMI was correlated with a slight increase in the likelihood of long COVID, which indicated that for each unit increase in BMI, the odds of developing long COVID increased by 3%. Asian or Asian British patients showed the highest odds of developing long COVID compared to other ethics. Former smokers were over 6.44 times more likely to develop long COVID compared to non-smokers. Comorbidities were strongly associated with long COVID, with patients having one or more comorbid conditions showing significantly higher odds.

Patients with ILDs were found to have a higher risk of developing long COVID. The study showed a significant association between Clauss fibrinogen, serum Na^+^ levels and the odds of long COVID; a one-unit increase in Clauss fibrinogen increased the risk by 22%, and a one-unit increase in sodium increased the risk by 7%. Patients who were fully vaccinated and received booster doses showed higher odds of developing long COVID, but this was insignificant.

The most common symptoms were fatigue (81%), shortness of breath (61%), dry/nonproductive cough (19%), chest pain, short-term memory issues, and memory difficulties were 12% each and anxiety (9.1%).

### Interpretation of Findings

These results emphasise the importance of a multifaceted approach in understanding long COVID, considering demographic, health, and behavioural factors to improve patient outcomes. Our findings emphasise the significance of age as a factor contributing to long COVID. Each additional year of age increased the odds of developing long COVID by 6.4%. This finding aligns with previous research indicating that older age increases susceptibility to long COVID ^42–47^. Increased age is associated with a higher likelihood of developing long COVID, a phenomenon that may be attributed to the deterioration of immune function that occurs with ageing ^44^. Also, it has been postulated that older patients have chronic conditions such as DM and heart and lung diseases, which could complicate recovery from COVID-19 and increase the risk of long COVID symptoms^48^. Studies have further suggested that the SARS–CoV–2 virus or its remnants could persist in the body for extended periods, potentially causing ongoing symptoms^49^. This persistence might be more pronounced in older patients due to their less robust immune response ^49^. Moreover, ageing is associated with increased levels of inflammation in the body. This chronic inflammation can exacerbate the effects of COVID-19 and contribute to prolonged symptoms ^48^.

The study showed an increase in long COVID risk associated with higher BMI. Studies have shown that obesity is associated with an increased risk of severe COVID-19 outcomes, which in turn correlates with a higher incidence of long COVID symptoms ^50,51^. The underlying mechanisms may involve systemic inflammation and impaired immune responses associated with obesity, which can prolong recovery and contribute to the persistence of symptoms ^52^.

Moreover, the interaction between age and BMI further complicates the risk landscape for long COVID. Research has indicated that younger individuals with higher BMI may face a greater risk of severe COVID-19 outcomes compared to older adults, where the relationship between BMI and disease severity appears to diminish ^53,54^. This suggests that while age is a critical factor, the impact of obesity on long COVID may be particularly pronounced in younger populations.

Ethnic minorities emerged as another significant factor, with Asian or Asian British patients showing the highest odds of developing long COVID. This aligns with findings from studies that suggest ethnic minorities, particularly South Asians, face higher risks for severe COVID-19 outcomes^55^. This observation may be attributed to genetic polymorphisms that affect the host’s immune responses to infections, including those caused by SARS-CoV-2 ^56^. The specific genetic profiles might predispose individuals to more severe or prolonged symptoms. The lack of significant associations observed for Black and White individuals contrasts with numerous studies indicating heightened risks within Black populations. This discrepancy highlights the variability that may arise across diverse demographic groups and research contexts ^56^.

Also, individuals with a history of smoking (former smokers) demonstrated over 15-fold increased odds of developing long COVID. This stark contrast between former and current smokers’ risks points to the long-term impacts of smoking history on respiratory and overall health status. These findings support WHO’s evidence linking smoking to severe COVID-19 and related complications, underlining the need for addressing smoking cessation and its lingering effects^57^. Smoking causes long-term damage to the lungs and respiratory system, which can impair the body’s ability to recover from infections like COVID-19 ^58^. Also, smoking weakens the immune system, making it harder for the body to fight off infections and recover fully. This weakened immune response can lead to prolonged symptoms^58^.

Comorbidities demonstrate a strong correlation with long COVID, where individuals with one or more coexisting conditions exhibit markedly higher odds of persistence in symptoms. This observation aligns with existing literature suggesting that the concurrent management of multiple health issues can exacerbate recovery trajectories following SARS-CoV-2 ^59^. Recent studies indicate that various chronic conditions, including diabetes and cardiovascular diseases, are linked to persistent inflammation. This chronic inflammatory state may intensify the detrimental effects of COVID-19 and play a role in the manifestation of long COVID symptoms ^59^. Pre-existing conditions can significantly compromise organ function, particularly in the lungs, heart, and kidneys. The presence of COVID-19 can exacerbate this damage, resulting in more severe manifestations of disease and extended symptomatology ^60^.

Furthermore, our study found that patients with ILDs were at high risk of developing long COVID, reinforcing the importance of managing these underlying conditions to mitigate long COVID risks. This is aligned with a study that revealed that ILDs cause scarring and inflammation of the lung tissue, significantly impairing lung function. This pre-existing damage makes it harder for the lungs to recover from the additional stress and damage caused by COVID-19 ^61^. Interstitial lung diseases (ILDs) are characterised by persistent inflammatory processes within the pulmonary interstitium. The pathophysiology of COVID-19 can intensify this inflammatory response, resulting in exacerbated clinical manifestations and prolonged respiratory symptoms.

The association between vaccination status and long COVID remains complex. Our findings indicate that fully vaccinated individuals with booster doses exhibit marginally increased odds of long-term COVID outcomes, albeit this is not significant. This, though, stands in contrast to existing literature suggesting that vaccination mitigates the risk of long-term symptoms. This inconsistency may arise from variations in study cohorts, the timing of vaccinations, and the emergence of different variants of concern, highlighting the need for further research to clarify these effects.

Among various biomarkers, Clauss fibrinogen was significantly associated with an increased risk of long COVID. Elevated Clauss fibrinogen levels, which indicate inflammation and coagulation, suggest a potential biomarker for susceptibility to prolonged COVID-19 symptoms. Fibrinogen is a key protein in the blood clotting process. During COVID-19 infection, the virus can cause abnormal clotting, leading to the formation of microclots.

These microclots can persist and contribute to long-term symptoms by obstructing blood flow and causing tissue damage^62^. Elevated levels of fibrinogen are also associated with increased inflammation. COVID-19 can trigger a strong inflammatory response, and high fibrinogen levels can exacerbate this inflammation, leading to prolonged symptoms^62^. Studies have shown that fibrinogen can form dense, resistant clots that are difficult for the body to break down. These clots can trap inflammatory molecules, contributing to ongoing symptoms such as fatigue, cognitive impairment, and respiratory issues ^63^. This finding aligns with emerging research on the role of inflammation in COVID-19 pathophysiology, offering a new avenue for predictive and preventive strategies.

In addition, serum sodium (Na^+^) concentration correlated with an increased risk of developing long COVID. The relationship between serum Na+ concentration and long COVID is an emerging area of research that highlights the complex interplay between electrolyte imbalances and the long-term sequelae of COVID-19. While specific studies directly linking serum sodium levels to long COVID are limited, several related findings provide insights into the broader context of how electrolyte disturbances, particularly sodium levels, may influence long COVID outcomes. A study found a link between low electrolyte levels and fatigue among long COVID sufferers, implying that maintaining proper electrolyte balance is key to symptom management ^64^. Another study reveals that systemic inflammation persists in COVID-19 survivors, potentially affecting electrolyte balance and contributing to long COVID symptoms^65^. The inflammatory milieu observed in long COVID patients may lead to alterations in renal function, impacting sodium retention and excretion, thereby influencing serum sodium levels^65^. Elevated IL-6 levels have been associated with persistent symptoms following COVID-19 infection, which may indirectly affect sodium levels through mechanisms such as increased vascular permeability and altered renal function ^66^. This suggests that the inflammatory response in long COVID could lead to dysregulation of sodium homeostasis, potentially resulting in hyponatremia, which has been observed in various inflammatory states ^66^.

The wide range of symptoms affecting multiple body systems is consistent with the diverse symptomatology reported in long COVID literature. Neurological manifestations, such as fatigue, memory issues, and dizziness, were commonly reported and aligned with the findings from previous studies ^67,68^. The significant prevalence of psychological symptoms such as anxiety and depression highlights the mental health challenges associated with long COVID findings that align with previous research in the field ^69^. Other studies also commonly reported respiratory and cardiovascular manifestations such as shortness of breath, dry cough, chest pain, and chest tightness.

## Findings Implications

The identification of key risk factors for long COVID, including age, Asian or Asian British ethnicity, history of smoking, comorbidities, and ILDs, establishes a critical basis for developing targeted interventions. Healthcare providers can leverage on these findings to prioritise monitoring and support efforts for high-risk populations, particularly older adults, ethnic minorities, and former smokers. Rehabilitation programmes and follow-up care can be tailored to address the broad spectrum of symptoms exhibited, which encompass neurological, respiratory, and psychological domains, thereby facilitating a holistic approach to patient management.

This study adds to the expanding body of literature on long COVID by validating known risk factors while identifying novel associations. The significant association of Clauss fibrinogen and Na^+^ as a biomarker highlights its potential for further investigation into the pathophysiological mechanisms associated with long COVID. A deeper understanding of these mechanisms could facilitate the development of predictive models and targeted therapeutic interventions, ultimately improving the management strategies for long COVID symptoms.

## Strengths and Limitations

One significant strength was the relatively large sample size of 627 participants, which provided substantial data for identifying patterns and associations related to long COVID. Using a diverse participant pool increases the generalisability of our results across different demographic groups. Another significant strength was the use of PSM as a statistical analysis method to reduce bias. This holistic perspective allows for a more thorough understanding of long COVID’s multifaceted nature, contributing valuable insights to the existing body of knowledge. However, limitations were acknowledged as a retrospective study. The study’s observational nature could only identify associations but not direct causal relationships. The study design limited our ability to track symptom(s) progression and changes over time, which could provide deeper insights into the long COVID.

## Conclusion

Our study has delineated pivotal risk factors linked to the development of long COVID, including advanced age, a specific ethnicity minority, a history of smoking, and the presence of underlying comorbidities, particularly interstitial lung diseases (ILDs). Furthermore, we have identified novel correlations, notably the significant role of Clauss fibrinogen and serum Na^+^ as potential biomarkers and the complex implications of vaccination status on outcomes. Long COVID’s extensive systemic effects are highlighted in the study, emphasising the need for a multidisciplinary approach to manage the diverse symptoms in affected individuals.

This research enhances our comprehension of long COVID and paves the way for further research. Further research is essential to clarify the pathophysiology of long COVID and develop effective preventative, diagnostic, and therapeutic approaches to improve health outcomes for those affected.

## Institution review board

The Health Research Authority (HRA), England, and Health and Care Research Wales (HCRW) approved the study with REC reference 23/HRA/1637.

## STROBE statement

The authors have read the STROBE guideline, and our report was prepared following the Strengthening the Reporting of Observational Studies in Epidemiology (STROBE) guidelines.

## Author contributions

Lanre P. Daodu was responsible for developing the study protocol, conducting the study, analysing the data, and drafting the manuscript. Yogini Raste, Francesca I.F. Arrigoni, and Judith E. Allgrove provided oversight and reviewed the study protocol and the manuscript. Reem Kayyali oversaw the study, reviewed the protocol and manuscript, and provided overall supervision.

## Data Availability

All data produced in the present study are available upon reasonable request to the authors.

